# “FGFR4 p.Gly388Arg Mutations in PBMCs of LAM Patients: A Potential Systemic Driver of Neoplastic-like Behaviour”

**DOI:** 10.1101/2024.11.05.24316663

**Authors:** Sinem Koc-Gunel, Amy L. Ryan, Melanie Winter, T.O.F. Wagner

## Abstract

Lymphangioleiomyomatosis (LAM) is a rare, progressive lung disease with neoplastic traits, primarily driven by mutations in the TSC2 gene, which lead to hyperactivation of the mTOR pathway and uncontrolled cell growth. However, additional mutations may contribute to disease progression. In this study, we investigate the potential role of co-mutations, focusing on the FGFR4 p.Gly388Arg polymorphism, which has previously been associated with aggressive cancer progression, as a possible co-driver in LAM. Peripheral blood mononuclear cells (PBMCs) were isolated from seven sporadic LAM patients and analyzed using Next-Generation Sequencing (NGS) to identify tumorigenic mutations. The FGFR4 p.Gly388Arg variant was identified in four patients, with allelic frequencies ranging from 49% to 99%. The highest frequency was observed in a patient with severe bullous lung disease, who ultimately required lung transplantation. Our analysis revealed a strong positive correlation (r = 0.85, p = 0.15) between FGFR4 allelic frequency and lung function decline (FEV1%), as well as a moderate positive correlation (r = 0.55, p = 0.20) between FGFR4 mutation status (presence vs. absence) and FEV1% decline. Although these correlations did not reach statistical significance, the trends suggest that FGFR4 mutations may contribute to disease progression in LAM. These findings indicate that FGFR4 mutations could play a role in the systemic nature of LAM, potentially exacerbating disease severity. Further research is needed to evaluate FGFR4 as a biomarker and therapeutic target in conjunction with mTOR inhibitors for the treatment of LAM.

## Introduction

Lymphangioleiomyomatosis (LAM) is a rare, progressive multisystem disorder characterized by the abnormal proliferation and phenotype alteration of smooth muscle-like cells surrounding blood vessels, bronchioles, and lymphatic vessels (1). This pathological process leads to cystic lung lesions, respiratory failure, and tumours in the kidneys and retroperitoneum. While traditionally considered a benign lung disease, the World Health Organization classifies LAM as a perivascular epithelioid tumour (PeComa) due to its neoplastic features. LAM cells have been found in various bodily fluids, including the lymphatic system, and the disease has recurred in case studies after lung transplantation, suggesting metastatic potential (2). The mechanism by which LAM cells spread across organs or enter the circulatory system remains unclear. Despite their relatively low proliferative index, LAM cells share several characteristics with cancer cells. Recent studies, including our own, have described activated αSMA-positive fibroblasts in LAM, which exhibit increased invasiveness compared to healthy fibroblasts (3). Additionally, lymphangiogenesis and extracellular matrix (ECM) remodelling in LAM, driven by growth factors like VEGF-D, VEGF-A, and FGF7, further resemble processes seen in cancer (4). The current understanding of sporadic LAM is linked to somatic mutations in sporadic LAM and germline mutations in LAM associated with Tuberous sclerosis index and loss of heterozygosity (LOH) in the TSC2 gene, and less frequently TSC1, resulting in disinhibition of the mTOR pathway and unchecked proliferation of smooth muscle-like cells as well as cytoskeletal alterations. In TSC-LAM, about a quarter of patients carry TSC1 mutations, though these are rarer in sporadic LAM. Mutations in both genes vary widely, with no specific hotspots, including missense, large deletions, and in-frame deletions (5,6). Treatment therefore currently focuses on targeting LAM effector cells with TSC mutations using rapamycin, an mTOR inhibitor. However, rapamycin does not reverse lung damage or fully halts disease progression, highlighting the need for alternative therapeutic strategies. Neoplastic diseases often involve both driver mutations, which directly promote tumor growth, and co-driver mutations, which further influence disease progression. In LAM, mutations in the TSC2 gene are recognized as the primary driver, activating the mTOR pathway. However, as in other cancers, additional mutations may contribute to disease progression. Mutations beyond TSC2, particularly in genes regulating cell proliferation and signaling, may critically influence LAM pathogenesis. In this study, we explored the presence of additional mutations, such as co-driver mutations, in LAM and evaluated their potential clinical significance.

## Methods

### Patient Collective

This study included seven sporadic LAM patients monitored at a large academic hospital in Germany over 8 years. Peripheral blood mononuclear cells (PBMCs) were isolated from each patient. Clinical data, including lung function measurements (FEV1%), patient age, and treatment regimens (such as the use of rapamycin), were collected. Written informed consent was obtained from all patients and the study was approved by the responsible Institutional Review Board and the Ethics Committee (STO-02-2017). LAM was diagnosed based on standard diagnostic criteria according to American Thoracic Society (ATS) guidelines, which include typical cystic lung lesions identified on high-resolution CT (HRCT) scans, and in some cases through additional screening of serum VEGF-D levels (above 800 pg/mL) as a biomarker for LAM. In cases where VEGF-D testing was not conducted, diagnosis was confirmed by a combination of clinical and radiological findings. For five out of seven patients, the diagnosis was additionally confirmed through lung biopsy in conjunction with the radiological findings. Patients underwent PFTs at various intervals, with the number of tests per year varying between individuals, depending on clinical need and follow-up schedules, over a follow-up period ranging from 1 to 8 years.

### PBMC Isolation and NGS Mutation Analysis

#### PBMC Isolation

PBMCs were isolated from whole blood with a standardized protocol using a Ficoll gradient.

#### DNA Extraction and Quantification

Genomic DNA was isolated as previously described (7). Briefly, genomic DNA was extracted using the Maxwell® RSC Instrument (Promega Corporation) with the Maxwell® RSC FFPE Plus DNA Kit (Promega Corporation), following the manufacturer’s protocol. DNA quantification was performed using the Qubit 4 Fluorometer (Invitrogen). For each sample, 20 ng of DNA was used as input for the library preparation.

#### Next-Generation Sequencing Using the Oncomine Comprehensive Assay

Targeted mutation analysis was performed using the Oncomine Comprehensive Assay v3 (Thermo Fisher Scientific), an NGS platform designed to detect tumorigenic mutations across a wide panel of cancer-related genes. The assay identifies single nucleotide variants (SNVs), insertions and deletions (INDELs), copy number variations (CNVs), and gene fusions. Libraries were prepared using the Ion AmpliSeq™ technology and 20 ng of DNA as input.

#### Library Preparation and Sequencing

DNA libraries were prepared according to the manufacturer’s protocol using the Ion AmpliSeq™ Library Kit 2.0. The libraries were pooled and subsequently sequenced on the Ion S5 System (Thermo Fisher Scientific). Raw sequencing data were processed using Torrent Suite™ software for primary analysis. Data analysis was further carried out using Ion Reporter™ software (version 5.12.0.0) with filter chains Oncomine Variants 5.10 and Oncomine Extended 5.12.

#### Data Analysis

Post-sequencing data analysis focused on identifying relevant mutations in the FGFR4 gene, particularly the p.Gly388Arg mutation. Other tumorigenic mutations in genes such as TSC2, BRCA2, and NOTCH3 were also evaluated. The results were analyzed using Ion Reporter™, with identified variants cross-referenced against public databases for clinical relevance. Quality control metrics for the sequencing data adhered to the platform’s cut-off values. The allelic frequency of each mutation was calculated, and data were validated by cross-referencing with control samples.

#### Lung Function Assessment

PFTs, including spirometry and body plethysmography, were performed using a body plethysmograph following the recommendations of the American Thoracic Society and the European Respiratory Society. The following parameters were measured: forced vital capacity (FVC), forced expiratory volume in 1 second (FEV1), FEV1/FVC ratio, residual volume (RV), residual volume/total lung capacity (RV/TLC), and diffusing capacity of the lungs for carbon monoxide (DLCO).

#### Correlation Analysis

To investigate the relationship between FGFR4 mutations and lung function decline in LAM patients, two variables were analyzed: FGFR4 mutation status and FGFR4 allelic frequency. Mutation status was coded as 1 for the presence of a mutation and 0 for its absence. FEV1% decline was used as a measure of lung function deterioration. Pearson’s correlation analysis was performed to assess the association between FGFR4 mutation status and FEV1% decline. A scatter plot was generated in R (version 2024.04.2+764) using the ggplot2 package, with FEV1% decline plotted on the y-axis and FGFR4 mutation status on the x-axis. A linear regression model was applied to visualize the relationship, and a fitted regression line was included in the plot. The association between FEV1% decline and FGFR4 allelic frequency was also analyzed using Pearson’s correlation. For this analysis, allelic frequency was treated as a continuous variable, and the ggplot2 package was used to create scatter plots and regression lines to visualize the relationship. All analyses adhered to standard statistical protocols, ensuring accurate data handling and interpretation.

## Results

Drawing on advances in liquid biopsy technology from the lung cancer field, we aimed to investigate whether LAM patients harbour detectable systemic mutations in circulating cells that may contribute to disease progression. To address this, we isolated PBMCs from seven LAM patients and performed NGS targeted mutation analysis using the Oncomine Comprehensive Assay v3 (Thermo Fisher Scientific). This high-throughput, NGS panel widely standartized for clinical settings assesses mutations in over 160 cancer-related genes, including key oncogenes, tumour suppressor genes, and genes associated with tumorigenesis pathways.

The clinical and genetic characteristics of the patients included in the study are included in **Table 1**. This is the first study to identify FGFR4 mutations in PBMCs of LAM patients, supporting systemic involvement in LAM progression. Our analysis revealed the FGFR4 p.Gly388Arg mutation in four of seven patients at chromosome 5:176520243, with allelic frequencies of 49%, 49%, 99%, and 51%. The highest allelic frequency (99%) was observed in STLAM1, a patient with pronounced bullous lung changes (**Fig. 1)**, occurrence of pneumothorax in past medical history, and lymphangioleiomyoma, treated with sirolimus. Another patient, STLAM2, had a 49% FGFR4 mutation frequency alongside a TSC2 mutation (56%) and exhibited cystic lung remodeling, pleural effusions, and angiomyolipomas (AML), also receiving sirolimus. Similar systemic features were observed in STLAM6, with past medical history of pleural effusions, chylothoraces, and abdominal lymph node involvement, who also carried the FGFR4 mutation (49%) but had no detectable TSC2 mutation. Lastly, STLAM3 had a 51% FGFR4 mutation frequency and exhibited cystic lung changes, though she was not receiving any treatment. In addition to FGFR4 mutations, we detected a TSC2 mutation (p.Ile1735HisfsTer40) at chr16:2138268 in STLAM2 with an allelic frequency of 56%, and a BRCA2 mutation (p.Ile2177MetfsTer13) at chr13:32915019 with a low allelic frequency of 4% in STLAM1. An additional mutation in NOTCH3 (p.Asp1086Ala) was found in STLAM2 with a low allelic frequency of 6.83%. Patients STLAM5, STLAM4, and STLAM7 did not show FGFR4 mutations, although they exhibit cystic lung disease with no detectable systemic TSC2 mutations.

**Table 1:** Genetic Characteristics of LAM Patients.

| Patient ID | STLAM1 | STLAM2 | STLAM3 | STLAM4 | STLAM5 | STLAM6 | STLAM7 |
| --- | --- | --- | --- | --- | --- | --- | --- |
| Age Range | 50-55 | 56-60 | 56-60 | 65-70 | 50-55 | 56-60 | 65-70 |
| Gender | Female | Female | Female | Female | Female | Female | Female |
| LAM Type | Sporadic | Sporadic | Sporadic | Sporadic | Sporadic | Sporadic | Sporadic |
| FGFR4 Mutation | Yes | Yes | Yes | No | No | Yes | No |
| FGFR4 Allelic Frequency (%) | 99 | 49 | 51 | N/A | N/A | 49 | N/A |
| FGFR4 Mutation Chromosome Locus | chr5:176520243 | chr5:176520243 | chr5:176520243 | N/A | N/A | chr5:176520243 | N/A |
| FGFR4 Mutation Type | SNP | SNP | SNP | N/A | N/A | SNP | N/A |
| Mutation Coverage | 1989,1992, 1959, 1995 | 1989, 1992, 1959, 1995 | 1989, 1992, 1959, 1995 | N/A | N/A | 1989, 1992, 1959, 1995 | N/A |
| TSC2 Mutation | No | Yes | No | No | Yes | No | No |
| TSC2 Allelic Frequency (%) | N/A | 56 | N/A | N/A | N/A | N/A | N/A |
| TSC2 Mutation Chromosome Locus | N/A | chr16:2138268 | N/A | N/A | N/A | N/A | N/A |
| TSC2 Mutation Type | N/A | INDEL | N/A | N/A | N/A | N/A | N/A |
| TSC2 Mutation Coverage | N/A | 1993 | N/A | N/A | N/A | N/A | N/A |
| Treatment | Sirolimus | Sirolimus | None | None | Sirolimus | Sirolimus | None |

**Fig. 1:**
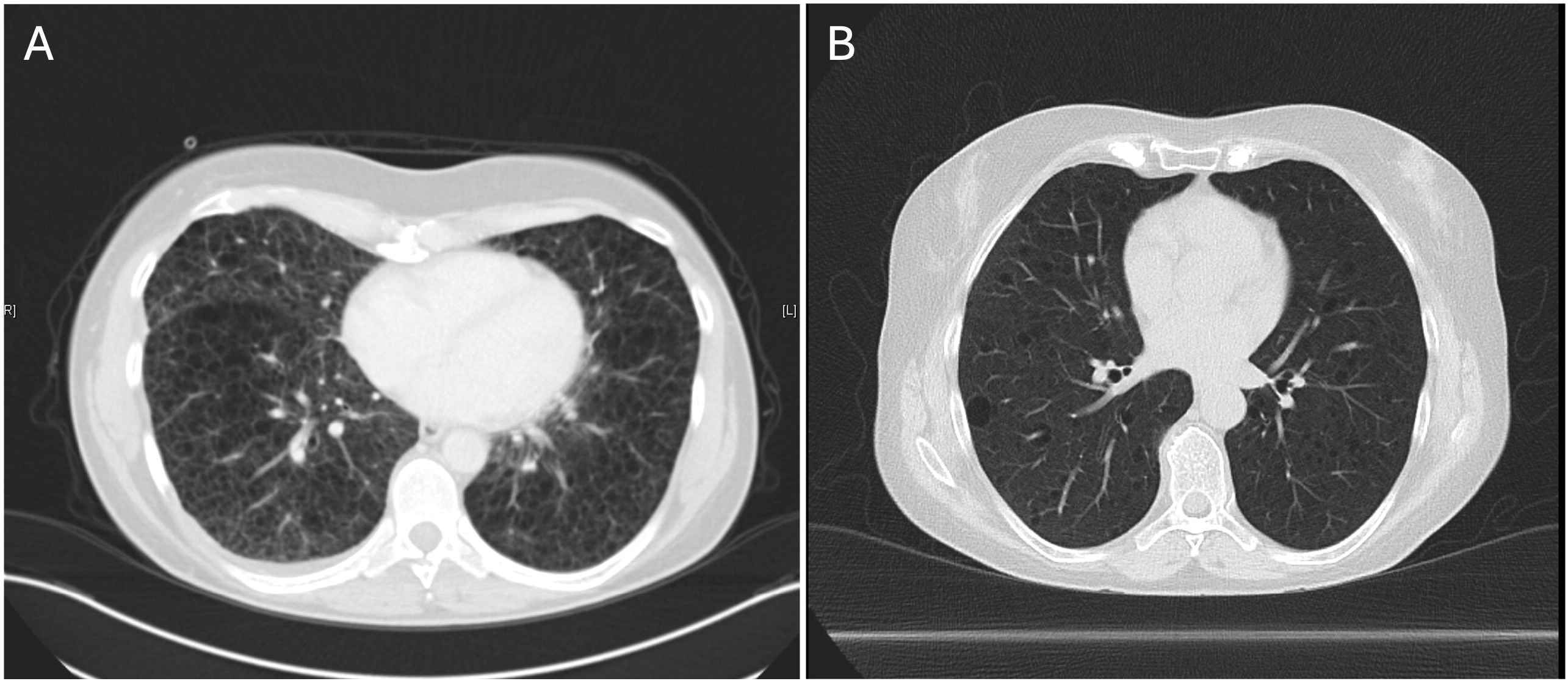
Representative lung CT-Scans of STLAM1 and STLAM7. (A) CT images from STLAM1, with a high allelic frequency (99%) of the FGFR4 p.Gly388Arg mutation, demonstrate advanced cystic destruction. (B) In contrast, STLAM7, with no FGFR4 p.Gly388Arg mutation, exhibits bilateral cysts of varying sizes alongside preserved lung areas.

To further investigate the relationship between FGFR4 mutations and disease progression, we analyzed the correlation between the allelic frequency of the FGFR4 p.Gly388Arg mutation and the rate of decline in lung function (FEV1%) over time. In total, we analyzed 52 pulmonary function tests (PFTs) from 7 patients. **Figure 2** shows the individual FEV1% progress for each patient over time. Pearson’s correlation analysis was performed to examine the association between FGFR4 allelic frequency and the rate of lung function decline. Our results revealed a strong positive correlation (r = 0.85), though the relationship did not reach statistical significance (p = 0.15) **(Fig.3)**. This finding suggests that higher allelic frequency of the FGFR4 mutation may be associated with a faster decline in lung function, potentially contributing to disease progression in LAM patients. Additionally, we analyzed the relationship between FGFR4 mutation status (presence vs. absence) and FEV1% decline. This analysis revealed a moderate positive correlation (r = 0.55, p = 0.20) between mutation status and lung function decline, suggesting that patients harboring FGFR4 mutations may experience a more pronounced decline in FEV1% compared to those without mutations **(Fig.4)**. Although the results are not statistically significant, the observed trends highlight the potential importance of FGFR4 mutations in disease progression. These findings warrant further investigation in larger studies to confirm their clinical significance.

**Fig. 2:**
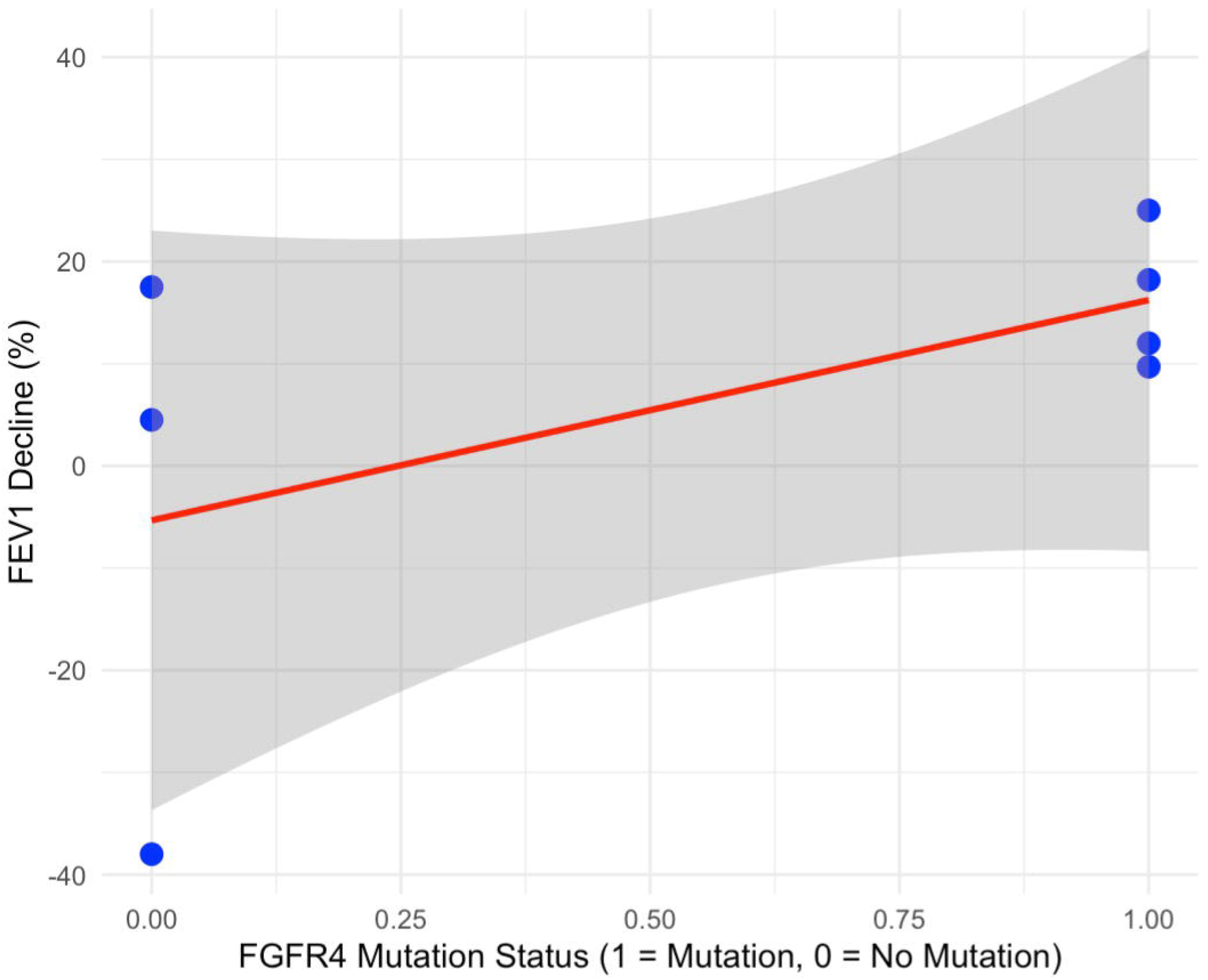
FEV1 (%) over time and FGFR4 mutation status for LAM patients. FEV1 (%) over time in LAM patients, with distinct colors representing individual patients. The figure shows a longitudinal assessment of lung function, highlighting variations in decline rates across patients with and without FGFR4 mutations.

**Fig. 3:**
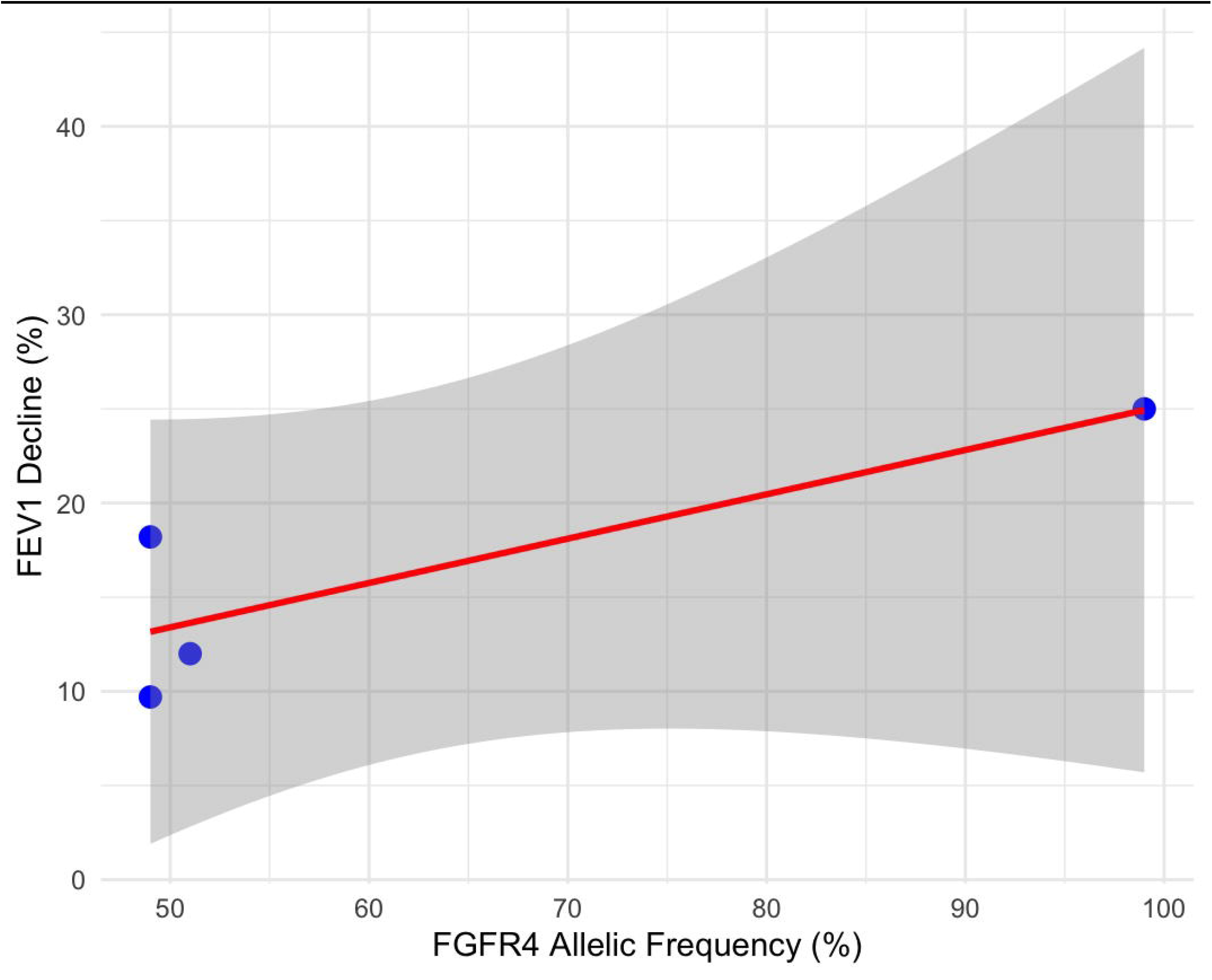
Correlation Between FGFR4 Allelic Frequency and FEV1% Decline in LAM Patients. The scatter plot shows the relationship between FGFR4 allelic frequency (%) and lung function decline (FEV1%) among LAM patients carrying the FGFR4 mutation. Each point corresponds to an individual patient, with the red regression line highlighting the observed trend. A strong positive correlation (r = 0.847, p = 0.153) suggests that higher FGFR4 allelic frequencies may be associated with accelerated lung function decline. The analysis was performed using Pearson’s correlation and a linear regression model.

**Fig. 4:**
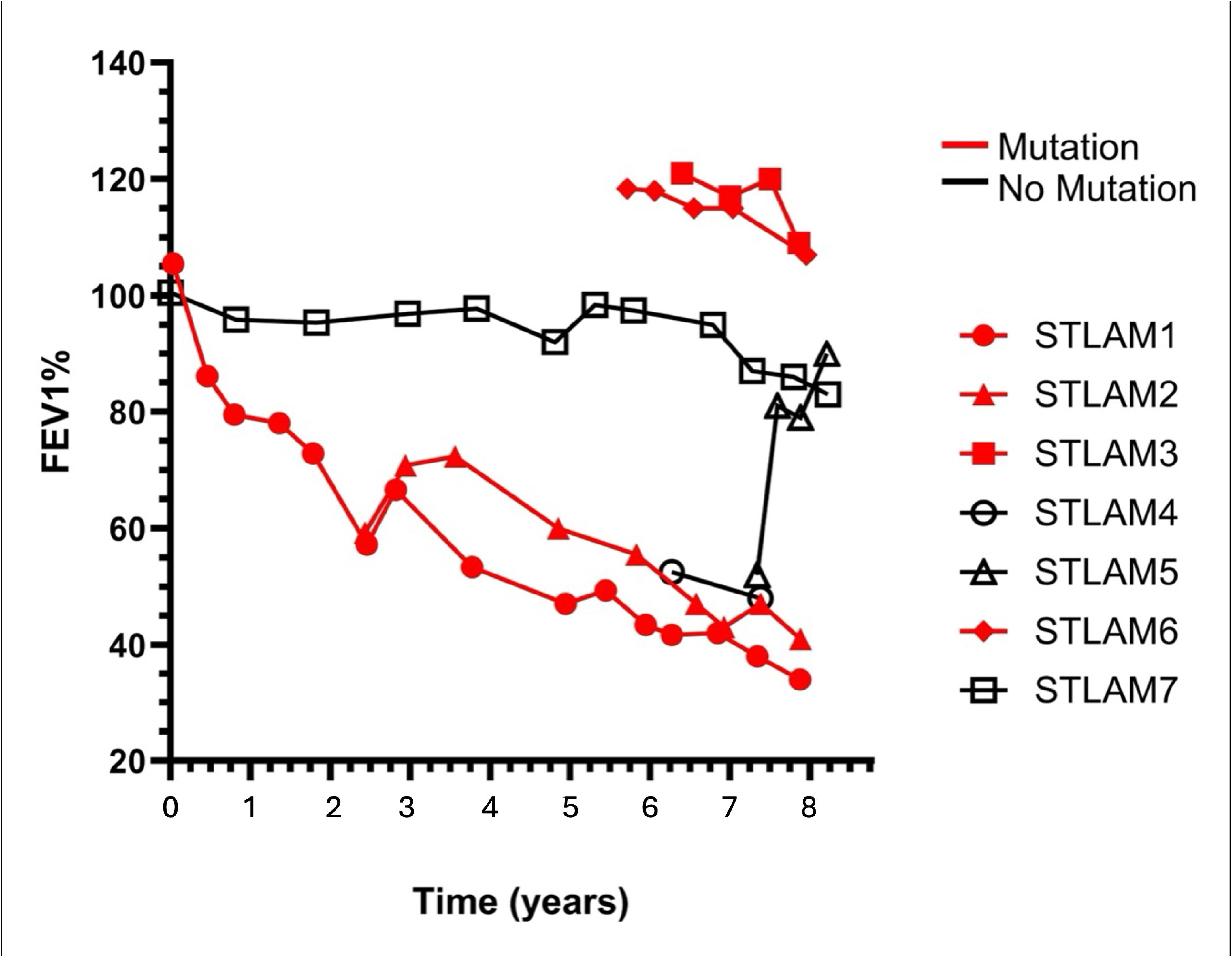
Correlation between FGFR4 mutation status and FEV1% decline in LAM patients. The scatter plot illustrates the relationship between FGFR4 mutation status (1 = mutation present, 0 = mutation absent) and the percentage decline in lung function (FEV1%) among LAM patients. Each point represents an individual patient, and the red regression line indicates the trend. A moderate positive correlation (r = 0.551, p = 0.200) suggests that patients with FGFR4 mutations may experience a greater decline in lung function compared to those without mutations. Data points were analyzed using Pearson’s correlation and a linear regression model.

## Discussion

Our study is the first to describe genetic alterations in PBMCs derived of LAM patients, which is suggestive of systemic involvement in LAM and reiterating it as a systemic neoplastic disease. Previous studies have established TSC2 mutations as the primary driver of LAM. However, our data introduces FGFR4 mutations in PBMCs as a potential co-driver, expanding the molecular landscape of LAM beyond the mTOR pathway. This aligns with research indicating that FGFR4 mutations, particularly the FGFR4 p.Gly388Arg variant, may negatively impact the prognosis of neoplastic diseases. A systematic review by Kim et al. demonstrated that FGFR4 p.Gly388Arg polymorphisms are associated with worse outcomes in cancer patients (8). Although LAM is typically a slow-progressing neoplasm, the discovery of this polymorphism in immune cells in peripheral blood may have several important implications.

First, it is important to note that while this variant has been identified at lower allelic frequencies in population studies, including a large study analyzing 16,179 controls, it is present in approximately 33% of the global population, with variability depending on ethnicity (9,10). In contrast, our findings show a significantly higher frequency in LAM patients. Despite the small sample size, we identified the FGFR4 p.Gly388Arg mutation in 4 out of 7 patients (56%), suggesting a potential role in LAM disease pathology that warrants further exploration. FGFR4 p.Gly388Arg has been described repeatedly in contributing to cell motility and lymph node metastasis in various types of cancers including but not limited to lung, breast, head, neck, gastric and colorectal cancer (10–17). A metaanalysis including 8555 subjects on the association of between the FGFR4 p.Gly388Arg polymorphism and cancer risk showed higher susceptibly to develop malignancies for this specific SNP (18). Two other metanalyses showed a poor prognosis for the same polymorphism in patients diagnosed with cancer (12,19). Studies looking at the downstream impact of the polymorphism show that a change the pGly388Arg polymorphism leads to an activated STAT pathway contributing to epithelial-to-mesenchymal transition (14,20). The SNP results in the substitution of glycine with arginine at codon 388 in the FGFR4 receptor and has been further described in vivo, in vitro, and in silico to lead to changes in the receptor domain as well as induce MAP kinase activation, likely contributing to increased cell proliferation, survival, and tumorigenesis (16,17). Given LAM’s neoplastic and metastatic characteristics, a subpopulation of LAM patients with the FGFR4 p.Gly388Arg mutation may be at risk for worse outcomes, suggesting that this gene polymorphism could be a key factor in disease progression. For example, patient STLAM1, who exhibited a 99% allelic frequency of this mutation, experienced more severe disease progression, including the development of bullous cysts in the lungs, which ultimately led to lung transplantation. This contrasts sharply with the clinical course of STLAM7, who did not carry the FGFR4 polymorphism and had a less aggressive disease trajectory during a similar observation period. This divergence in clinical outcomes underscores the potential role of FGFR4 p.Gly388Arg in exacerbating disease severity and accelerating lung function decline in LAM patients, indicating that this polymorphism could be an important factor in disease progression and therapeutic response. Our sample size is limited, and these findings require further validation through larger studies and in vitro experiments. Future research should explore the potential cross-talk between FGFR4 and mTOR pathways. FGFR4 mutations could enhance the effects of TSC2 mutations by activating downstream pathways such as MAPK/ERK and PI3K/AKT, both of which are known to stimulate mTOR signaling. This crosstalk between FGFR4 and mTOR pathways may promote unchecked cell proliferation and survival in LAM, contributing to disease progression. These insights suggest that targeting FGFR4 in addition to mTOR could be a potential therapeutic strategy. Importantly, the detection of FGFR4 mutations in PBMCs, which include T-lymphocytes, B-lymphocytes and natural killer (NK) cells populations, suggests a potential role for immune dysregulation in LAM. Although the role of FGFR4 mutations in immune cells in LAM is not well understood, research in other neoplasms suggests that such mutations in immune cells can contribute to disease progression. Finally, in this study a TSC2 mutation was identified in patient STLAM2, who also had an FGFR4 mutation. Notably, STLAM2 did not exhibit typical clinical features of tuberous sclerosis, which is caused by germline TSC2 mutations, suggesting a more nuanced interplay between TSC2 and FGFR4 mutations in LAM pathogenesis. The presence of the TSC2 mutation in PBMCs raises the possibility of systemic involvement, potentially acting alongside FGFR4 mutations to amplify pro-fibrotic processes or support unchecked proliferation of LAM cells. These findings underscore the need for further investigation into how immune cell dysfunction and the interaction between FGFR4 and TSC2 mutations may exacerbate lung fibrosis and drive LAM progression. Our findings introduce FGFR4 as a potential co-driver in LAM progression, suggesting that FGFR4-targeted therapies may offer new therapeutic avenues, particularly in patients with both FGFR4 mutations. Further studies with larger cohorts are needed to validate these findings, as well as in vitro and in vivo studies to explore the mechanistic interplay between FGFR4 and mTOR signaling pathways.

## Data Availability

All data produced in the present study are available upon reasonable request to the authors.

## Data Availability Statement

The genomic datasets analyzed for this study can be found in the **KocG-nelLAB_FGFR4LAM** GitHub repository. The repository includes.bam.bai files for each sequencing alignment. Additional data or information can be provided upon reasonable request.

